# Laboratory verification of new commercial lateral flow assays for Cryptococcal antigen (CrAg) detection against the predicate IMMY LFA in a reference laboratory in South Africa

**DOI:** 10.1101/2022.03.02.22271757

**Authors:** Lindi-Marie Coetzee, Deborah Kim Glencross

## Abstract

**Background:** Reflex Cryptococcal antigen (CrAg) testing in HIV-positive patients is done routinely at 47 laboratories in South Africa on samples with a confirmed CD4 count <100 cells/μl, using the IMMY Lateral Flow Assay (LFA) as the standardized predicate method.

**Objective:** This study aimed to verify the diagnostic performance of newer CrAg LFA assays against the predicate method.

**Methods:** Remnant CD4 samples collected between February and June 2019, with confirmed predicate LFA CrAg results, were retested on settled plasma with the (i) IMMY CrAg semi-quantitative (SQ) LFA; (ii) Bio-Rad RDT CryptoPS SQ; and (iii) Dynamiker CrAg SQ assays, within 24 hours of predicate testing. Sensitivity/ specificity analyses were conducted comparing predicate versus the newer assays, with McNemar’s test’s p-values reported for comparative results (p values <0.05 significant). Positivity grading was noted for the IMMY SQ and Bio-Rad assays.

**Results:** Of the 254 samples tested, 228 had comparative CrAg results across all assays. The predicate method reported 85 CrAg positive (37.2%), compared to between 35.08 and 37.28% for the Bio-Rad, IMMY SQ and Dynamiker assays. The IMMY CrAg SQ grading (+1 to +5) showed 67% of CrAg positive results had a grading ≥3, indicative of higher CrAg concentration (infection severity). False-negative results across all assays were <2%, with sensitivity >95% for all. False-positive results were highest for the Dynamiker LFA (14%) with a specificity of 77% (p=0.001). IMMY SQ and Bio-Rad assays specificities exceeded 90% (p=0.6 and 0.12). Internal quality control showed 100% accuracy for all assays.

**Conclusion:** Performance verification of newer CrAg LFA assays under typical laboratory conditions varied, with best results by IMMY SQ and Bio-Rad. The high burden of HIV and cryptococcal disease in South Africa requires high specificity and - sensitivity (>90%) to prevent unnecessary treatment/hospitalization. The added value of positivity grading for patient management needs confirmation.

## Introduction

Cryptococcal meningitis (CM) is a highly infectious disease caused by *Cryptococcus neoformans*, with a high mortality rate, particularly among HIV-positive patients in developing countries such as South Africa [1-3]. Following the inclusion of cryptococcal antigen (CrAg) screening in the 2016 World Health Organization (WHO) HIV guidelines [4], and subsequent inclusion in the South African HIV guidelines [5], a CrAg reflex screening pilot program was launched at selected National Health Laboratory Service (NHLS) CD4 laboratories in South Africa [6, 7]. This program was subsequently extended nationally in June 2017 to 47 CD4 laboratories, where remnant blood samples from HIV-positive patients with a confirmed CD4 count <100cells/μl (severely immune-compromised) are identified by the laboratory information system (LIS) for reflex CrAg testing [8-10]. The data collected for the program has been used extensively to report on the prevalence of CrAg in South Africa [8, 9, 11], the cost-effectiveness of the local program [12, 13] and the high burden of patients living with HIV with a CD4 count <100 cells/μl (immune-compromised) [8, 9]. Approximately 10% of all CD4 samples tested annually (~230 000/annum) receive a reflex CrAg test with a national CrAg positivity rate of 6.2% [8, 9].

All reflexed CrAg testing is performed using the lateral flow assay (LFA) from IMMY (Immuno-Mycologics, IMMY, Norman, OK, USA) [14]. The IMMY LFA was the only commercially available assay at the time of the national roll-out and was recommended in the WHO HIV guidelines from 2018 [15]. Several studies described the performance of this assay against the ELISA and latex agglutination assays on serum, plasma, cerebrospinal fluid and urine [14, 16-30]. Since 2018, additional CrAg LFA products have been developed and introduced worldwide and became available in South Africa [31-33].

This study set out to verify the diagnostic performance of the newer CrAg LFA assays against the predicate method (IMMY LFA) for accuracy, sensitivity and specificity under typical laboratory conditions. The high burden of HIV and cryptococcal disease in South Africa requires an assay with high specificity and sensitivity (>90%) to prevent unnecessary treatment/hospitalization. This will enable the accurate distinction of positive and negative results to ensure appropriate early initiation of antifungal therapy to reduce mortality.

## Materials and Methods

### Ethics clearance

Ethics clearance was obtained from the University of the Witwatersrand (M1706108, approved for 5 years from 13/07/2017 to 07/2022). No patient consent was required as random remnant blood samples were used as per ethics clearance after laboratory predicate results were reported. The study results were for assay verification purposes only. No patient information was available to either the testing staff or the authors of this paper.

### CrAg Testing methods

All reagents and test consumables were supplied by local diagnostic suppliers, with training provided for each assay for two medical technologists. The assays verified in this study are CE/IVD approved (Conforms to European Union Requirements/in vitro diagnostics). The manufacturer instructions defined in the package insert were used for reagents storage, quality control (positive and negative internal controls), testing (sample volume, incubation time), interpretation of results, detection limitations and result reporting. All safety precautions were adhered to as part of good laboratory practice (GLP). Results were reported as positive, negative, or invalid (no control line visible), with concentration ratings noted for specific assays as per their package inserts (IMMY SQ and Bio-Rad). Samples with faint positive lines were verified by a senior staff member or retested if the result was inconclusive. All CrAg testing and retesting were done on settled plasma, with no titrations performed as this is not the practice for CrAg reflex screening.

#### Laboratory predicate method

The predicate method is the LFA by IMMY (Immuno-Mycologics, Norman, Oklahoma, United States) [34]. This assay is an immunochromatographic test for the qualitative and semi-quantitative detection of the capsular polysaccharide antigen of *Cryptococcus* species in plasma and serum, using specimen wicking and gold-conjugated anti-CrAg antibodies. Currently, the predicate method is used only as a qualitative assay for reflex testing. Samples were prepared according to the manufacturer’s instructions and the national internal standard operating procedure.

#### Newer generation CrAg LFA methods

This IMMY CrAg SQ dipstick sandwich immunochromatographic assay is a newer commercial IMMY CrAg dipstick (Immuno-Mycologics, Norman, Oklahoma, United States) [33]. The principle is similar to the original IMMY CrAg LFA assay, with the exception that there is one control line and two test lines (T1 and T2) on a wicking strip [33, 35]. Positive results will always have a control line, with either T1 and/or T2 lines visible. The intensity (concentration) of CrAg binding is interpreted from a scoring scale of 1+ to 5+ as per package insert.

The Bio-Rad RDT CryptoPS (Biosynex CryptoPS) assay is a single-use rapid semi-quantitative CrAg strip test (BIOSYNEX S.A., Illkirch–Graffenstaden, France) [36]. Sample and diluent are dispensed into the well and test lines (T1 and T2) will form depending on the concentration in the specimen [31, 36]. T1 represents concentrations lower than 25ng/ml with T2 detecting concentrations up to 2.5μg/ml. Dynamiker CrAg LFA (Dynamiker Technology, Tianjin Eco-City, Tianjin, China) [32] is an immunochromatographic test using conjugated Cryptococcus antibodies to gold particles. CrAg will form complexes that appear as a visible line, while the free antibodies bind to form a control line [32].

### Patient samples and study settings

Remnant samples collected in Ethylenediaminetetraacetic acid (EDTA) submitted for CD4 testing at the Charlotte Maxeke Academic Hospital (CMJAH) and Tambo Memorial laboratories between February and May 2019, with a confirmed count of <100cells/μl, received onsite reflex CrAg testing using the IMMY CrAg LFA predicate method. These laboratories are accredited by the South African National Accreditation System (SANAS), adhere to good laboratory practice (GLP) and International Organization for Standardization (ISO/ICE 15189:2014) guidelines [37]. In addition, they are enrolled in the local external quality assessment (EQA) [38]. Patient management was based on the predicate CrAg result reported.

Reagents for 300 tests were provided from local service providers for IMMY and Dynamiker (only 280 tests available from Bio-Rad). Before comparative testing commenced, four tests per assay were reserved for training and ten tests for possible duplicate/repeat testing. Daily quality-control tests at two levels (negative and positive) were done per assay/testing day (n=32 tests/assay in total) (Fig 1).

**Figure 1:**
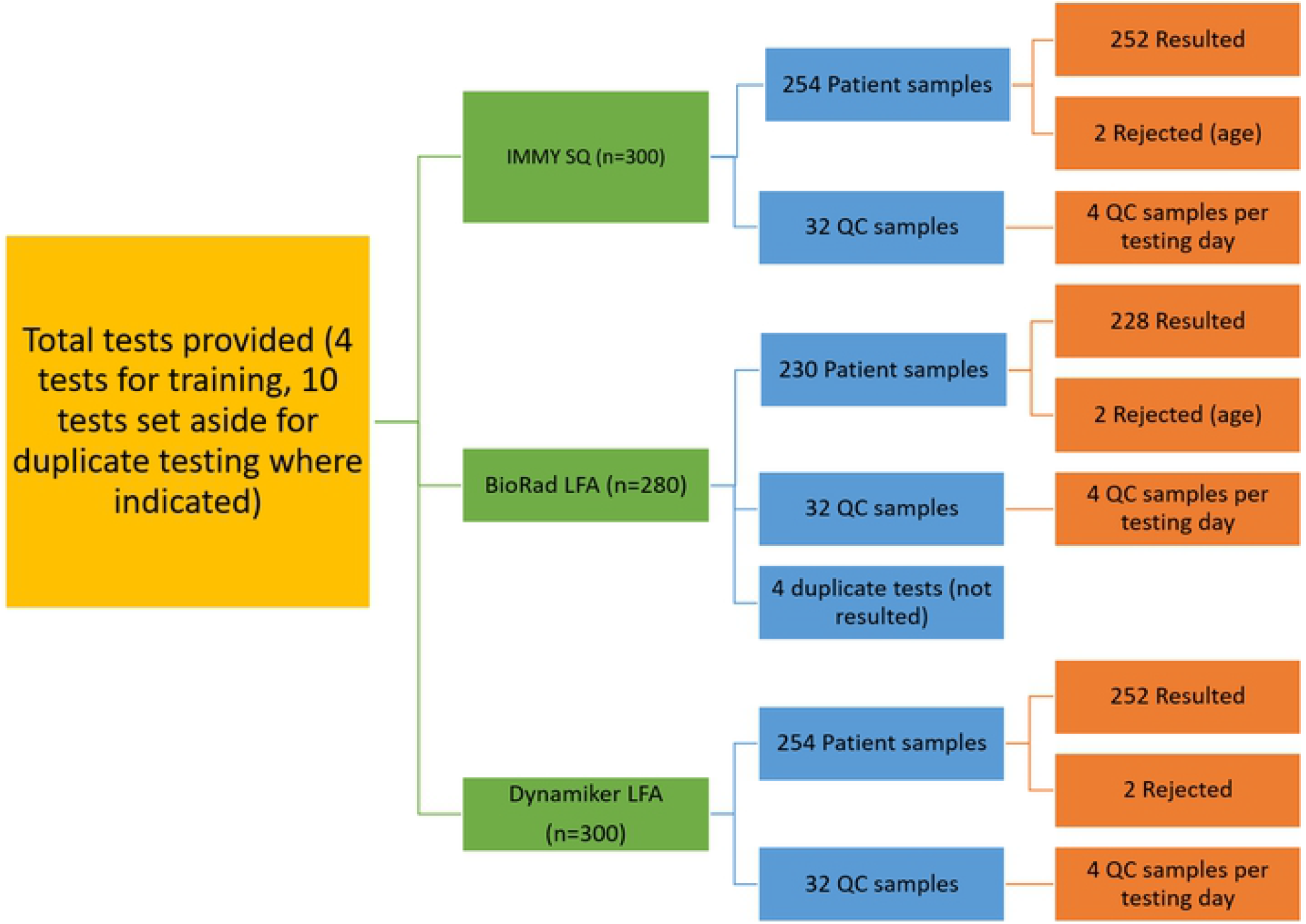
Summary of verification samples tested: Patient samples tested across three new CrAg LFA assays, including quality control (QC), duplicate testing and rejections, using the available tests provided by suppliers. (Authors own work).

Following predicate testing, samples were delivered to the CMJAH CD4 research unit for re-testing within 24 hours. The project coordinator collated CrAg results in and Excel spreadsheet and batched available samples for blind testing. The batch size was restricted to a maximum of 25 samples (n=75 tests across 3 assays) per day (≤ 3 hours per testing day per trainer) and included all positive samples collected from the testing laboratories on the testing day with random negative samples. Results for the IMMY CrAg SQ LFA [33], Bio-Rad RDT CryptoPS Assay (Biosynex CryptoPS) [31, 36], and Dynamiker CrAg LFA assays [32] were recorded on printed worklists against anonymized sample ID numbers and collated by the project coordinator into the project Excel file.

The verification criteria for sensitivity and specificity levels were set at >90% to correlate with the current IMMY CrAg LFA predicate method. The Standards for Reporting of Diagnostic Accuracy Studies (STARD) statement checklist was used for transparency of result reporting [39, 40].

### Statistical analysis

Statistical analysis and the graphic display were done with GraphPad Prism Software version 7 (GraphPad Software, San Diego, United States). Assay performance statistics included: sensitivity, specificity, positive predictive value (PPV), negative predictive value (NPV) and accuracy. These parameters were calculated using the predicate IMMY CrAg LFA as the reference method. The MedCalc software and online calculator were used for this purpose [41], with McNemar’s test’s for paired nominal data used to assess significance between the predicate and each new CrAg assay, with p-values calculated (p<0.05 regarded as significant) [42].

## Results

Assay-specific positive and negative controls were analyzed with every batch of samples retested with 100% accuracy for all assays over the test period. Of the 10 tests per assay set aside for duplicate or re-testing, only 4 repeat tests were performed with the Bio-Rad assay. Repeat test results were not reported as the outcome did not change from the original result observed.

### Performance of reference and test methods

#### Reference methods results

In total, 254 patient samples were tested using the IMMY LFA SQ and Dynamiker SQ assays. Only 230 samples could, however, be retested using the Bio-Rad assay due to import challenges and the available timeline of the project. With 2 samples excluded due to receipt >72 hours after predicate testing, only 228 samples could be used for comparison across the three new assays (Fig 1). Of these 228 results, the predicate LFA CrAg reference method identified 143 CrAg negative (62.72%) and 85 CrAg positive results (Table 1).

**Table 1:** Summary of all CrAg assay results.

| IMMY Predicate CrAg LFA as Reference | IMMY SQ | BioRad | Dynamiker |
| --- | --- | --- | --- |
| Positive number (% of total) | 84 (36.84) | 81 (35.52) | 85 (37.28) |
| Negative number (% of total) | 140 (61.40) | 132 (57.89) | 111 (48.68) |
| False-positive number (% of total) | 3 (1.31) | 11 (4.82) | 32 (14.03) |
| False-negative number (% of total) | 1 (0.43) | 4 (1.75) | 0 (0.0) |
| p-value (Mc Nemar test) | 0.617 | 0.123 | <0.001*** |
| **TOTAL** | **228** | **228** | **228** |

The total test number and number of positive and negative samples were recorded as well as the number of false-positive and false-negative samples for each CrAg assay compared to the predicate CrAg LFA assay as reference method (n=85 CrAg positive and 143 CrAg negative samples). Data is reported as test results recorded and the percentage of total tests (n=228), with a p-value of the McNemar's test's for paired nominal data comparison (<0.05 statistically significant).

Of all samples tested, 183/228 (80.3%) showed equivalent results across all testing assays versus predicate; 102 negative (71.3% of total negative samples) and 81 positive samples (95.3% of total positive samples). The remaining 45/184 samples showed discrepant results for one or more assays against the reported predicate result (41 and 4 reported a negative and positive result respectively) (Fig 2).

**Figure 2:**
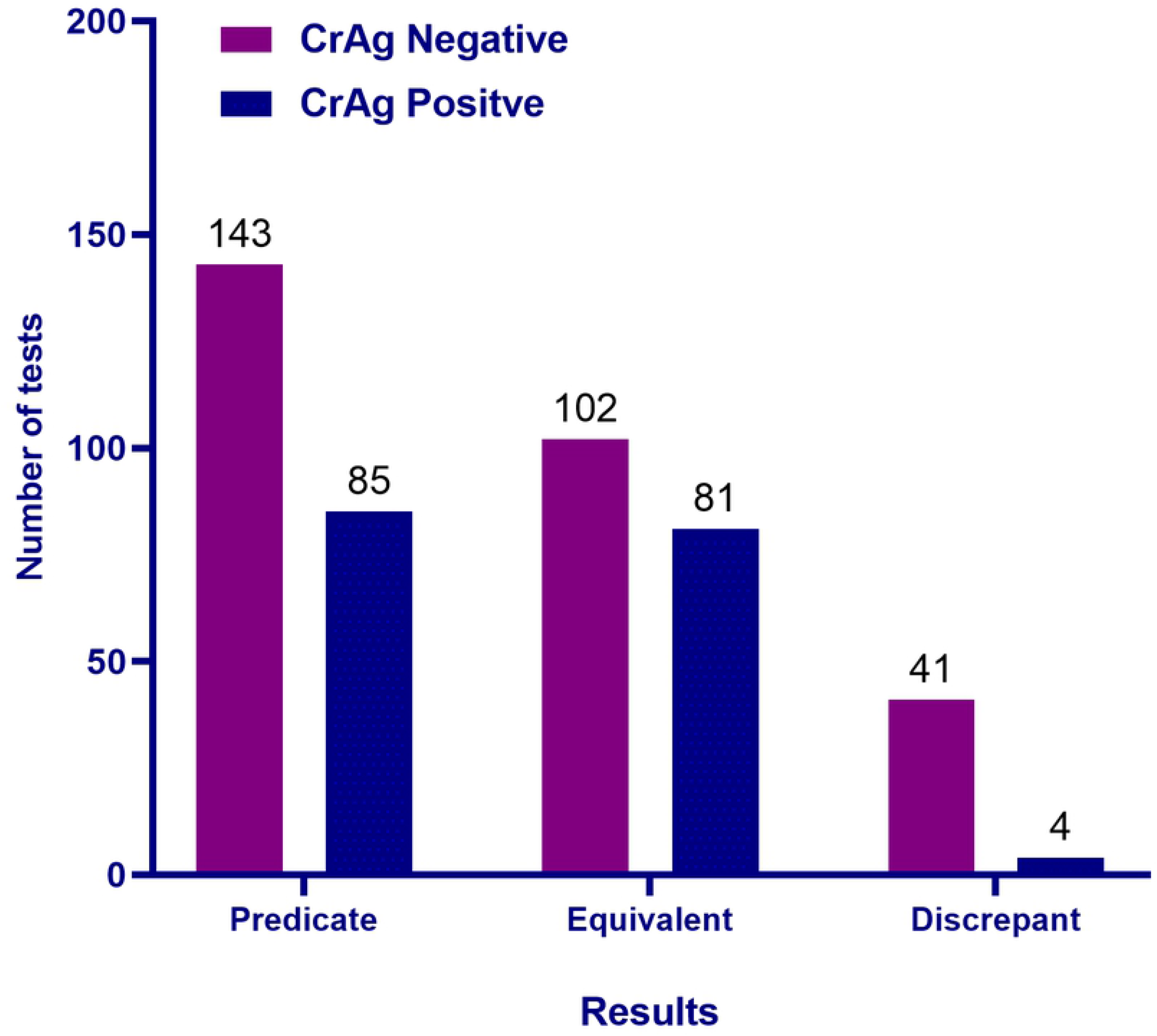
CrAg test results compared to the predicate LFA method: A summary of CrAg test results by the predicate method, indicating the number of results that were equivalent across the four LFA assays versus the numbers of discrepant results, where one or more assays had opposite results to the predicate result reported. (Purple bars represent CrAg negative results and blue bars CrAg positive results),

### Newer commercial CrAg LFA test assay results

Excellent agreement was noted for the number/percentage positive samples identified by the IMMY SQ, Bio-Rad and Dynamiker assays against the predicate method. The agreement for negative CrAg results, however, showed greater variability with the Bio-Rad and Dynamiker assays. False-positive results were observed with all assay comparisons to the predicate reference method used and ranged from 3 to 32 (1.31 to 14.03%) of the 228 samples tested, while false-negative results (compared to the reference method) of <2% were reported across the test assays (Table 1). The highest number of discordant results was reported for the Dynamiker assay, against the reference predicate method (p-value <0.001, McNemar’s test’s test). Of the 32 samples classified as false-positive, 11 had very faint positive bands, while 21 samples had clear positive bands as reported by two independent observers. Further analysis of Dynamiker assay performance excluding the 11 samples with faint positive results, only managed to reduce the percentage of false-positive samples by 3%, i.e. false positivity rates remained greater than 9% (data not shown).

IMMY SQ results showed an excellent correlation with the predicate method (Table 1; McNemar’s test’s test p=0.62). Of the 87 reported positive CrAg results, 20 recorded a 1+ reading (including the 3 false-positives compared to the predicate), with 20 a 2+, 28 a 3+. 18 a 4+ and only one a 5+ result, confirming that 67.85% of samples had a score of 3+ or more, i.e. an elevated concentration of CrAg associated with a high burden of disease. With Bio-Rad (McNemar’s test’s test p=0.12), of the reported 92 positive samples, 51 had a T1 result, of which 50 (98.03%) correlated with an IMMY SQ of a1+ or 2+ result, while 41 Bio-Rad tests had a T2 result, of which 37 (90.24%) correlated with an IMMY SQ results of ≥3+.

### Sensitivity and Specificity analysis

Sensitivity and specificity analyses were done, using the predicate IMMY CrAg LFA as the reference method on 228 samples (Table 2). Specificity was greater than 95% for IMMY SQ assay, while 92.36% and 77.62% were reported for the Bio-Rad and Dynamiker assays respectively. Sensitivity ranged from 95.24% to 100% for the Bio-Rad and Dynamiker assays. An accuracy of 85.95% was reported for Dynamiker, compared to >90% reported for IMMY SQ and Bio-Rad assays.

**Table 2:** Summary of specificity and sensitivity analyses.

|  | Sensitivity and Specificity parameters as percentage (95% CI) | IMMY CrAg SQ LFA | Bio-Rad CrAg LFA | Dynamiker CrAg LFA |
| --- | --- | --- | --- | --- |
| **PREDICATE IMMY CrAg LFA** | Specificity | 97.90 (93.99-99.57) | 92.36 (86.74-96.13) | 77.62 (69.90-84.16) |
|  | Sensitivity | 98.82 (93.32-99.97) | 95.24 (88.25-96.65) | 100 (95.75-100.00) |
|  | Positive Predictive Value (PPV) | 96.55 (90.13-98.85) | 87.91 (80.44-92.78) | 72.65 (66.19-78.28) |
|  | Negative Predictive Value (NPV) | 99.29 (95.23-99.9) | 97.08 (92.73-98.86) | 100 |
|  | Accuracy | 98.25 (95.57-99.52) | 93.42 (89.38-96.27) | 85.96 (80.77-90.20) |

Each of the newer CrAg assays was assessed against the predicate IMMY CrAg LFA. All results are reported as a percentage with the 95% confidence interval in brackets. Data were collected from February to June 2019 at two testing laboratories in South Africa.

## Discussion

This study set out to verify the diagnostic performance of the newer CrAg LFA assays against the predicate method (IMMY LFA) for accuracy, sensitivity and specificity under typical laboratory conditions.

Of the newer commercial CrAg assays, IMMY SQ faired best, with sensitivity and specificity exceeding 95%, while Bio-Rad results were also acceptable with sensitivity and specificity greater than 90%. Similar acceptable outcomes have been reported for the IMMY SQ assay by Tenforde et al. and Temfack et al [43, 44].

The lower sensitivity of between 78-88% and specificity of more than 90% [43-46] have been reported for the Bio-Rad CryptoPS assay (also marketed as Biosynex CryptoPS) as well as lower specificity in samples with a low fungal burden (missed positivity) in these samples [43, 44] The verification results of this current South African comparison, confirmed the slightly lower sensitivity and specificity of the Bio-Rad CryptoPS assay, however still within local acceptable criteria of sensitivity and specificity of >90%. A possible explanation previously reported mentioned the differences described in the limit of detection between IMMY LFA (5ng/ml) vs. Bio-Rad (25ng/ml) [46].

Our local study reported acceptable sensitivity of more than 98% for the Dynamiker CrAg assay, but specificity rates of 77% (vs. predicate LFA). The Dynamiker assay reported the highest number of false-positive results, mainly due to weak positive bands reported by two independent observers. Even with these removed from all calculations, the false positivity rate remained at ~10%. Similar high false-positive results were reported by Kwizera et al, where they showed comparable sensitivity with IMMY LFA, but poor specificity in serum and plasma samples [35]. This may in part be due to the lower detection limit of Dynamiker (1.25ng/ml vs. 1.75 ng/ml for IMMY LFA). A small study (n=25) published in 2018, reported that Dynamiker had a brighter intensity of a positive result than comparative results with the IMMY LFA, and the authors suggested that a faint positive result with Dynamiker should be reported as CrAg negative with this assay [47]. This is, however, impractical in a typical laboratory setting where only one assay kit is used and laboratory staff is trained to report all positive results (faint or clear) as such. No verification testing is done routinely in the NHLS CrAg reflex program and comparative testing may only be available in research facilities for specific projects. The observed higher positivity could be attributed to the differences in methods compared to IMMY and other LFA strips, in that there is no diluent used. A comparative study on cryopreserved serum samples (n=162) reported acceptable specificity of 89% of the Dynamiker assay versus IMMY LFA [48].Their study had only a small number of positive samples (n=14; 8.6% of total tests), but reported an equivalent number of false-positive results (n=15; 9.3% of total tests), which they contribute to the lower detection threshold of the Dynamiker assay.

All findings of our study were shared with the local service provider and subsequently, a refined version of the assay was provided for testing. Unfortunately, the issue of faint false-positive results persisted (data not shown)

Our study showed that the performance of the IMMY SQ and Bio-Rad assays were comparable to IMMY LFA results with both sensitivity and specificity greater than 90% These results confirm earlier reported verification results with this CrAg assay [43, 44, 46], where there was a good correlation with LFA titer results, where the grading from 1+ to 5+ showed correlation with titers [43, 44, 46]. This was further confirmed in a study on cerebrospinal fluid (CSF), where increasing IMMY SQ grades were associated with greater LFA titer and quantitative culture results/colony forming units (CFU/ml) [49]. The risk stratification offered by the IMMY SQ and Bio-Rad assays may be important from a clinical perspective to identify the severity of meningitis and the infiltration of the central nervous system (CNS) associated with elevated CrAg concentrations [43, 44, 46]. Patients with a grade of ≥3 may have a higher risk of poor outcomes, as reported by Tadeo et al, 2021 [49].

## Limitations

This verification study of the diagnostic performance of newer LFA CrAg test methods was done on typical laboratory samples, tested by qualified technologists. In-house unpublished reproducibility verifications performed on fresh EDTA samples over time using the IMMY LFA method showed reproducible results up to 48 hours on samples kept at room temperature. Assay recommendations indicate testing within 24 hours with samples stored at room temperature or up to 48 hours if refrigerated [32-34, 36]. Manufacturers need to take into consideration the time delay from sample collection to testing to ensure this does not affect the outcome of the test results, i.e. ensure the robustness of their assay to produce a reliable result even on samples older than 24 hours. Although the LFA technology is ideal for point-of-care testing at a clinic level, this would delay patient testing while waiting for a confirmation of a CD4 count, with most patients typically only returning for test results within 7-14 days [15].

Operator feedback highlighted extra preparation steps needed with some assays and expressed challenges with result interpretation where more than two bands were present, i.e. particularly with assays like Bio-Rad and IMMY SQ with multiple lines to read manually, even though all tests were read independently by two LFA trained members of staff.

Typical laboratory challenges with CrAg LFA testing include mislabeling and transcription errors to the worksheet and onto the LIS. Automated cassette readers could be used to ensure direct transfer of the strip result to the LIS. Some strip readers were tested locally, though the challenges were either it making use of an intermediary program (like Excel) for result reporting (no direct interface with laboratory information system), or single sample reading of results that may not necessarily reduce turn-around-time of CrAg result reporting or hands-on time by operators.

The positivity percentages reported here do not represent the incidence of CrAg in South Africa [50, 51] as samples were collected from two facilities for assay performance specifically. Due to fairly low positivity rates at these facilities, all positive samples available were retested. Care was taken to include as many positive samples for statistical comparisons between assays, within the limitations of suitable samples, available reagents and time of operators to conduct testing.

## Conclusion

The local verification of performance of newer commercial CrAg assays is necessary to confirm accurate result reporting as there is inevitable variability between assays. Additional information provided by some assays like the Bio-Rad and IMMY SQ i.e. the intensity of antigen detected should be further investigated to assess its value to clinicians in deciding on whether a lumbar puncture is needed or a CNS infiltration is suspected. The relevance of this added information may, however, depend on the local treatment guidelines i.e. in South Africa all CrAg positive patients will get a lumber puncture currently [15, 52] and cost-effectiveness of graded assays, especially in large national CrAg screening programs.

## Data Availability

All relevant data are within the manuscript and the Supporting Information file (S1).

## Acknowledgements

The authors would like to thank the trainers (Neo Mokone and Sithembile Mojalefa) of the National Priority Program (NPP) for testing samples.

## References

1. Pongsai P, Atamasirikul K, Sungkanuparph S. The role of serum cryptococcal antigen screening for the early diagnosis of cryptococcosis in HIV-infected patients with different ranges of CD4 cell counts. J Infect. 2010;60(6):474–7 https://doi.org/http://doi.org/10.1016/j.jinf.2010.03.015.

2. Jarvis JN, Boulle A, Loyse A, Bicanic T, Rebe K, Williams A, et al. High ongoing burden of cryptococcal disease in Africa despite antiretroviral rollout. AIDS. 2009;23(9):1 https://doi.org/10.1097/QAD.0b013e32832bc0fc

3. Parker BJ, Wannemuehler KA, Marston BJ N.P. G, Pappas PG, Chiller TM. Estimation of the current global burden of cryptococcal meningitis among persons living with HIV/AIDS. AIDS. 2009;23(4):5 https://doi.org/10.1097/QAD.0b013e328322ffac

4. World Health Organization. Consolidated guidelines on the use of antiretroviral drugs for treating and preventing HIV infection Recommendations for a public health approach [Guideline]. Geneva, Switzerland 2016, Accessed: 15 November 2019. Available from: https://www.who.int/hiv/pub/arv/arv-2016/en/.

5. Govender NP, Meintjes G, Mangena P, Nel J, Potgieter S, Reddy D, et al. Southern African HIV Clinicians Society guideline for the prevention, diagnosis and management of cryptococcal disease among HIV-infected persons: 2019 update. South Afr J HIV Med. 2019;20(1):1030 https://doi.org/10.4102/sajhivmed.v20i1.1030

6. Coetzee LM, Cassim N, Glencross DK, Rapid scale-up of reflexed Cryptococcal antigen (CrAg) screening across a CD4 laboratory network in South Africa. International Aids Society (IAS); 2017, 23-26 July; Paris, France, https://www.iasociety.org/Web/WebContent/File/IAS2017_conference_report.pdf.

7. Mokone G, Mojalefa S, Coetzee L, Glencross DK, Implementation of a National Cryptococcal antigen (CrAg) reflexed screening programme in South Africa. Society of Medical Laboratory Science of South Africa (SMLTSA); 2017, 19-21 May 2017; Durban, South Africa

8. Cassim N, Coetzee LM, Govender NP, Glencross DK. District and sub-district analysis of cryptococcal antigenaemia prevalence and specimen positivity in KwaZulu-Natal, South Africa. Afr J Lab Med. 2018;7(1):757 https://doi.org/10.4102/ajlm.v7i1.757

9. Coetzee LM, Cassim N, Sriruttan C, Mhlanga M, Govender NP, Glencross DK. Cryptococcal antigen positivity combined with the percentage of HIV-seropositive samples with CD4 counts <100 cells/ul identifies districts in South Africa with advanced burden of disease. PLoS One. 2018;13(6):e0198993 https://doi.org/10.1371/journal.pone.0198993

10. Govender NP, Glencross DK. National coverage of reflex cryptococcal antigen screening: A milestone achievement in the care of persons with advanced HIV disease. S Afr Med J. 2018;108(7):534–5 https://doi.org/10.7196/SAMJ.2018.v108i7.13094

11. Greene G, Lawrence DS, Jordan A, Chiller T, Jarvis JN. Cryptococcal meningitis: a review of cryptococcal antigen screening programs in Africa. Expert Rev Anti Infect Ther. 2020:1–12 https://doi.org/10.1080/14787210.2020.1785871

12. Cassim N, Coetzee LM, Schnippel K, Glencross DK. Estimating the cost-per-result of a national reflexed Cryptococcal antigenaemia screening program: Forecasting the impact of potential HIV guideline changes and treatment goals. PLoS One. 2017;12(8):e0182154 https://doi.org/10.1371/journal.pone.0182154

13. Cassim N, Schnippel K, Coetzee LM, Glencross DK. Establishing a cost-per-result of laboratory-based, reflex Cryptococcal antigenaemia screening (CrAg) in HIV+ patients with CD4 counts less than 100 cells/ul using a Lateral Flow Assay (LFA) at a typical busy CD4 laboratory in South Africa. PLoS One. 2017;12(2):e0171675 https://doi.org/10.1371/journal.pone.0171675

14. Kozel TR, Bauman SK. CrAg lateral flow assay for cryptococcosis. Expert Opin Med Diagn. 2012;6(3):245–51 https://doi.org/10.1517/17530059.2012.681300

15. World Health Organization. Guidelines for the diagnosis, prevention and management of cryptococcal disease in HIV-infected adults, adolescents and children [Guideline]. Geneva, Switzerland 2018, Accessed: 15 November 2019. Available from: https://www.who.int/hiv/pub/guidelines/cryptococcal-disease/en/.

16. Jarvis JN, Percival A, Bauman S, Pelfrey J, Meintjes G, Williams GN, et al. Evaluation of a novel point-of-care cryptococcal antigen test on serum, plasma, and urine from patients with HIV-associated cryptococcal meningitis. Clin Infect Dis. 2011;53(10):1019–23 https://doi.org/https://doi.org/10.1093/cid/cir613

17. Lindsley MD, Mekha N, Baggett HC, Surinthong Y, Autthateinchai R, Sawatwong P, et al. Evaluation of a newly developed lateral flow immunoassay for the diagnosis of cryptococcosis. Clin Infect Dis. 2011;53(4):321–5 https://doi.org/10.1093/cid/cir379

18. Binnicker MJ, Jespersen DJ, Bestrom JE, Rollins LO. Comparison of four assays for the detection of cryptococcal antigen. Clin Vaccine Immunol. 2012;19(12):1988–90 https://doi.org/10.1128/CVI.00446-12

19. Gates-Hollingsworth MA, Kozel TR. Serotype sensitivity of a lateral flow immunoassay for cryptococcal antigen. Clin Vaccine Immunol. 2013;20(4):634–5 https://doi.org/10.1128/CVI.00732-12

20. Hansen J, Slechta ES, Gates-Hollingsworth MA, Neary B, Barker AP, Bauman S, et al. Large-scale evaluation of the immuno-mycologics lateral flow and enzyme-linked immunoassays for detection of cryptococcal antigen in serum and cerebrospinal fluid. Clin Vaccine Immunol. 2013;20(1):52–5 https://doi.org/10.1128/CVI.00536-12

21. Vijayan T, Chiller T, Klausner JD. Sensitivity and specificity of a new cryptococcal antigen lateral flow assay in serum and cerebrospinal fluid. MLO Med Lab Obs. 2013;45(3):16, 8, 20

22. Kabanda T, Siedner MJ, Klausner JD, Muzoora C, Boulware DR. Point-of-care diagnosis and prognostication of cryptococcal meningitis with the cryptococcal antigen lateral flow assay on cerebrospinal fluid. Clin Infect Dis. 2014;58(1):113–6 https://doi.org/10.1093/cid/cit641

23. Lourens A, Jarvis JN, Meintjes G, Samuel CM. Rapid diagnosis of cryptococcal meningitis by use of lateral flow assay on cerebrospinal fluid samples: influence of the high-dose “hook” effect. J Clin Microbiol. 2014;52(12):4172–5 https://doi.org/10.1128/JCM.01683-14

24. Magambo KA, Kalluvya SE, Kapoor SW, Seni J, Chofle AA, Fitzgerald DW, et al. Utility of urine and serum lateral flow assays to determine the prevalence and predictors of cryptococcal antigenemia in HIV-positive outpatients beginning antiretroviral therapy in Mwanza, Tanzania. J Int AIDS Soc. 2014;17:19040 https://doi.org/10.7448/IAS.17.1.19040

25. Huang HR, Fan LC, Rajbanshi B, Xu JF. Evaluation of a new cryptococcal antigen lateral flow immunoassay in serum, cerebrospinal fluid and urine for the diagnosis of cryptococcosis: a meta-analysis and systematic review. PLoS One. 2015;10(5):e0127117 https://doi.org/10.1371/journal.pone.0127117

26. Chen M, Zhou J, Li J, Li M, Sun J, Fang WJ, et al. Evaluation of five conventional and molecular approaches for diagnosis of cryptococcal meningitis in non-HIV-infected patients. Mycoses. 2016;59(8):494–502 https://doi.org/10.1111/myc.12497

27. Nalintya E, Kiggundu R, Meya D. Evolution of Cryptococcal Antigen Testing: What is new? Curr Fungal Infect Rep. 2016;10(2):62–7 https://doi.org/10.1007/s12281-016-0256-3

28. Tang MW, Clemons KV, Katzenstein DA, Stevens DA. The cryptococcal antigen lateral flow assay: A point-of-care diagnostic at an opportune time. Crit Rev Microbiol. 2016;42(4):634–42 https://doi.org/10.3109/1040841X.2014.982509

29. Caceres DH, Zuluaga A, Tabares AM, Chiller T, Gonzalez A, Gomez BL. Evaluation of a Cryptococcal antigen Lateral Flow Assay in serum and cerebrospinal fluid for rapid diagnosis of cryptococcosis in Colombia. Revista do Instituto de Medicina Tropical de Sao Paulo. 2017;59:e76 https://doi.org/10.1590/s1678-9946201759076

30. Moodley K, Coetzee L, Glencross DK. Testing platforms for early detection of Cryptococcal Antigeneamia in high volume CD4 testing laboratories in South Africa. 8th INTEREST Workshop; 5-9 May; Lusaka, Zambia 2014. https://www.researchgate.net/publication/273577376_Testing_Platforms_for_Early_Detection_of_Cryptococcal_Antigenaemia_in_High_Volume_CD4_Testing_Laboratories_in_SouthAfrica

31. BioRad. RDT Crypto PS Assay 2017, Accessed: 20 October 2020. Available from: http://www.diagnostics-bio-rad.com/wp-content/uploads/2017/01/RDT-CryptoPS-Assay.pdf.

32. Biotechnology D. Dynamiker Cryptococcal Antigen Lateral Flow Assay (LFA) [Catalogue No.: DNK-1411-1]. London, England: Wellkang Ltd; 2017, Accessed: 9 October 2020. Available from: https://www.rafer.es/sites/default/files/pack_insert-dnk-cryptococcal_antigen_lfa.pdf.

33. IMMY. CrAgSQ Lateral Flow Assay: For the Detection of Cryptococcal antigen. Norman, OK, USA: IMMY; 2019.

34. IMMY. CrAg® LFA: CRYPTOCOCCAL ANTIGEN Norman, OK, USA: IMMY; 2019, Accessed: 2 October 2020. Available from: https://www.immy.com/crag.

35. Kiiza TK, Nimwesiga A, Skipper CP, Kwizera R, Apeduno L, Okirwoth M, et al., Diagnostic performance of a semi-quantitative Point of Care assay for Cryptococcis. CROI 2020; 2020, 8-11 March; Boston, MA, USA,https://www.croiconference.org/abstract/diagnostic-performance-of-a-semiquantitative-point-of-care-assay-for-cryptococcosis/.

36. Biosynex. BIOSYNEX® CryptoPS: For the semi-quantitative detection of Cryptococcus sp. antigens in whole blood, serum, plasma and CSF [Package insert 112001]. ILLKIRCH-GRAFFENSTADEN - France: Biosynex SA; 2018, Accessed: 9 October 2020. Available from: https://www.biosynex.com/flyers/pro/mycologie/en/cryptops.pdf.

37. South African National Accreditation System (SANAS). International and Regional Recognition [Regulatory guidelines]. Pretoria, South Africa: SANAS; 2020, Accessed: 12 October 2020. Available from: https://www.sanas.co.za/pages/index.aspx?page=international-regional-recognition.

38. National Health Laboratory Service (NHLS). Proficiency testing Johannesburg, South Africa 2021, Accessed: 29 April 2021. Available from: https://www.nhls.ac.za/quality-assurance/proficiency-testing/.

39. Cohen JF, Korevaar DA, Altman DG, Bruns DE, Gatsonis CA, Hooft L, et al. STARD 2015 guidelines for reporting diagnostic accuracy studies: explanation and elaboration. BMJ Open. 2016;6(11):e012799 https://doi.org/10.1136/bmjopen-2016-012799

40. Bossuyt PM, Reitsma JB, Bruns DE, Gatsonis CA, Glasziou PP, Irwig L, et al. STARD 2015: An Updated List of Essential Items for Reporting Diagnostic Accuracy Studies. Radiology. 2015;277(3):826–32 https://doi.org/10.1148/radiol.2015151516

41. MedCalc Software. Diagnostic test evaluation calculator: MedCalc Software Ltd, ; 2015, Accessed: 8 November 2021. Available from: https://www.medcalc.org/calc/diagnostic_test.php.

42. Trajman A, Luiz RR. McNemar chi2 test revisited: comparing sensitivity and specificity of diagnostic examinations. Scand J Clin Lab Invest. 2008;68(1):77–80 https://doi.org/10.1080/00365510701666031

43. Temfack E, Kouanfack C, Mossiang L, Loyse A, Fonkoua MC, Molloy SF, et al. Cryptococcal Antigen Screening in Asymptomatic HIV-Infected Antiretroviral Naive Patients in Cameroon and Evaluation of the New Semi-Quantitative Biosynex CryptoPS Test. Front Microbiol. 2018;9:409 https://doi.org/10.3389/fmicb.2018.00409

44. Tenforde MW, Boyer-Chammard T, Muthoga C, Tawe L, Milton T, Rulaganyang I, et al. Diagnostic accuracy of the Biosynex CryptoPS cryptococcal antigen semi-quantitative lateral flow assay in patients with advanced HIV disease. J Clin Microbiol. 2020 https://doi.org/10.1128/JCM.02307-20

45. Rajasingham R, Wake RM, Beyene T, Katende A, Letang E, Boulware DR. Cryptococcal Meningitis Diagnostics and Screening in the Era of Point-of-Care Laboratory Testing. J Clin Microbiol. 2019;57(1) https://doi.org/10.1128/JCM.01238-18

46. Skipper C, Tadeo K, Martyn E, Nalintya E, Rajasingham R, Meya DB, et al. Evaluation of Serum Cryptococcal Antigen Testing Using Two Novel Semiquantitative Lateral Flow Assays in Persons with Cryptococcal Antigenemia. J Clin Microbiol. 2020;58(58) https://doi.org/10.1128/JCM.02046-19

47. Bjornsdottir MK, Mahler SS, Raluca D, Arendrup MC, Evaluation of the new Dynamiker cryptotoccal antigen lateral flow assay (LFA) in comparison with IMMY LFA and Meridian latex agglutination test. 28th European Congress of Clinical Microbiology and Infectious Diseases (ECCMID) 2018, 21-24 April; Madrid, Spain, file:///C:/Users/lindi.coetzee/Documents/My%20Documents/2021/Papers/LFA%20review%201/P1197_abstract.pdf.

48. Noguera MC, Escandon P, Rodriguez J, Parody A, Camargo L. Comparison of two commercial tests (Immy vs. Dynamiker) for cryptococcal capsular antigen. Rev Soc Bras Med Trop. 2021;54:e03072021 https://doi.org/10.1590/0037-8682-0307-2021

49. Tadeo KK, Nimwesiga A, Kwizera R, Apeduno L, Martyn E, Okirwoth M, et al. Evaluation of the Diagnostic Performance of a Semiquantitative Cryptococcal Antigen Point-of-Care Assay among HIV-Infected Persons with Cryptococcal Meningitis. J Clin Microbiol. 2021;59(8):e0086021 https://doi.org/10.1128/JCM.00860-21

50. Coetzee LM, Cassim N, Glencross DK. Analysis of HIV disease burden by calculating the percentages of patients with CD4 counts <100 cells/microL across 52 districts reveals hot spots for intensified commitment to programmatic support. S Afr Med J. 2017;107(6):507–13 https://doi.org/10.7196/SAMJ.2017.v107i6.11311

51. Coetzee LM, Cassim N, Sriruttan C, Mhlanga M, Govender NP, Glencross DK. Cryptococcal antigen positivity combined with the percentage of HIV-seropositive samples with CD4 counts <100 cells/mul identifies districts in South Africa with advanced burden of disease. PLoS One. 2018;13(6):e0198993 https://doi.org/10.1371/journal.pone.0198993

52. Loyse A, Burry J, Cohn J, Ford N, Chiller T, Ribeiro I, et al. Leave no one behind: response to new evidence and guidelines for the management of cryptococcal meningitis in low-income and middle-income countries. Lancet Infect Dis. 2019;19(4):e143–e7 https://doi.org/10.1016/s1473-3099(18)30493-6

